# Ambient-Temperature Extraction-Free One-Pot CRISPR Detection of *Mycobacterium tuberculosis*

**DOI:** 10.64898/2026.07.21.26358604

**Authors:** Jipei Liao, Yun Su, Feng Jiang

## Abstract

**Background:** Current molecular diagnostics for *Mycobacterium tuberculosis* (MTB) require complex nucleic acid extraction procedures and laboratory infrastructure, limiting their use as point-of-care (POC) tests in resource-limited settings. To address these barriers, we developed an extraction-free one-pot CRISPR assay for rapid detection of MTB directly from minimally processed sputum specimens.

**Methods:** The assay integrates ambient-temperature chemical lysis, recombinase polymerase amplification, and CRISPR-Cas12a detection within a single closed-tube workflow. A conserved region of the MTB-specific *IS6110* insertion sequence was targeted for detection. Analytical performance was evaluated using serially diluted MTB genomic DNA standards, followed by clinical validation using 100 archived sputum specimens, including 50 MTB-positive samples and 50 MTB-negative controls.

**Results:** The extraction-free assay detected MTB genomic DNA within 30 minutes and achieved an analytical limit of detection of 100 copies per reaction. In clinical validation, the assay correctly identified 48 of 50 MTB-positive specimens and 49 of 50 MTB-negative specimens, yielding 96.0% sensitivity and 98.0% specificity.

**Conclusions:** This study demonstrates the feasibility of extraction-free one-pot CRISPR-Cas12a detection of MTB directly from sputum specimens. By eliminating conventional nucleic acid extraction while maintaining high analytical sensitivity and diagnostic performance, the platform may facilitate future development of rapid molecular diagnostics for POC and resource-limited settings.

## INTRODUCTION

Tuberculosis (TB), caused by *Mycobacterium tuberculosis* (MTB), remains a major global health threat, causing more than 10 million new cases and over one million deaths annually worldwide [1, 2]. Early and accurate diagnosis is essential for effective patient management, interruption of disease transmission, and implementation of public health control measures[3]. Current diagnostic approaches include sputum smear microscopy, mycobacterial culture, nucleic acid amplification tests, and radiographic evaluation [1, 4]. While culture remains the diagnostic reference standard, it is labor-intensive and requires prolonged incubation times [1]. Sputum smear microscopy is rapid and inexpensive but lacks sensitivity, particularly in patients with low bacterial burden [5]. Molecular assays such as GeneXpert provide improved sensitivity and specificity for tuberculosis diagnosis [6]. However, their dependence on specialized instrumentation, proprietary cartridges, and reliable supply chains can restrict implementation in decentralized and resource-limited settings [6].

Clustered Regularly Interspaced Short Palindromic Repeats (CRISPR)-associated (Cas) systems have emerged as powerful tools for nucleic acid detection because of their high analytical sensitivity and sequence specificity [7]. CRISPR-Cas12a-based diagnostics utilize guide RNA-mediated target recognition followed by collateral cleavage of reporter molecules, enabling rapid and sensitive detection of pathogen-derived nucleic acids [8]. Previous studies, including our own, have shown that the broad applicability of CRISPR-based platforms for detecting circulating tumor DNA, respiratory viruses, bacterial pathogens, and host immune biomarkers, including cytokines [9–16]. Recently, several CRISPR-based assays have been developed for detection of MTB DNA in clinical specimens[17]. These studies have demonstrated promising analytical performance; however, most workflows continue to rely on conventional nucleic acid extraction procedures, multiple processing steps, and laboratory-based infrastructure [17–29].

To address these challenges, we developed an extraction-free, one-pot RPA-CRISPR-Cas12a assay enabling rapid MTB detection directly from minimally processed sputum using an ambient-temperature, closed-tube workflow. The assay demonstrated excellent analytical and clinical performance, supporting its potential for point-of-care (POC) diagnosis in resource-limited settings.

## 2. MATERIALS AND METHODS

### 2.1. Overall Research Design

An extraction-free one-pot CRISPR-Cas12a assay was developed for rapid detection of MTB directly from minimally processed sputum specimens and evaluated for its analytical and clinical performance (Figure 1). The assay combines ambient-temperature chemical lysis with closed-tube recombinase polymerase amplification (RPA) and CRISPR-Cas12a detection in a streamlined sample-to-answer workflow. Analytical performance was first evaluated using defined MTB DNA samples, followed by clinical validation using archived sputum specimens from MTB-positive patients and MTB-negative controls. The overall workflow was designed to enable rapid molecular detection of MTB while minimizing sample handling and contamination risk.

**Figure 1.**
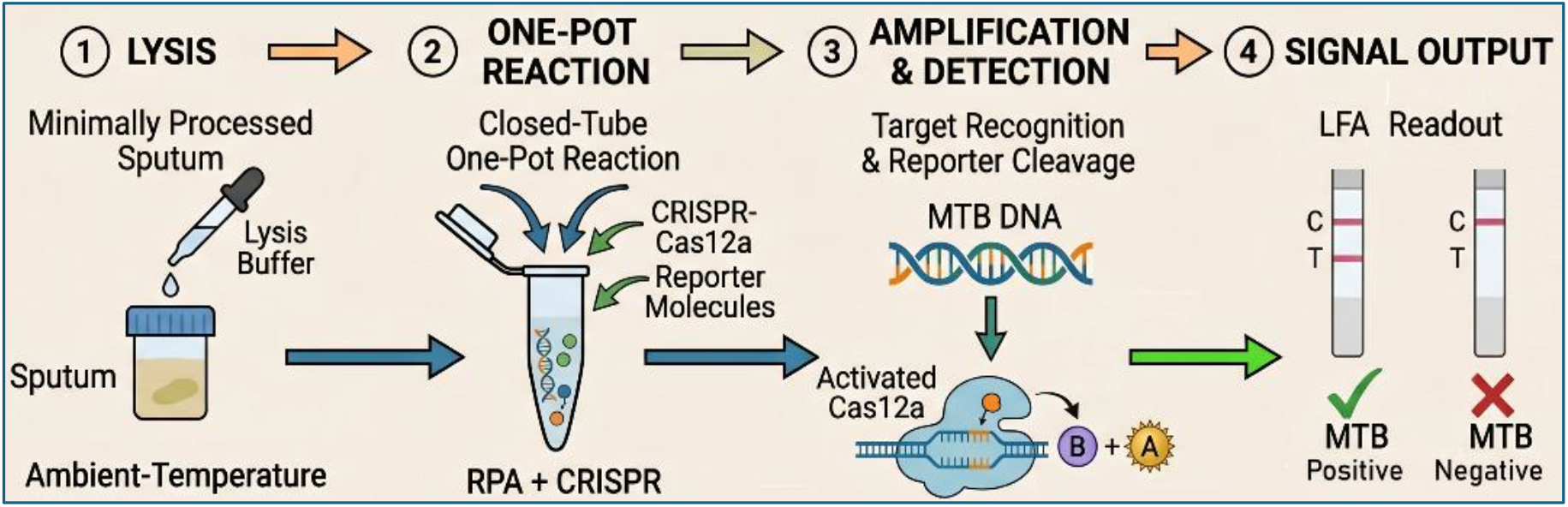
Schematic workflow of the extraction-free one-pot CRISPR-Cas12a assay for detection of MTB from minimally processed sputum. Sputum specimens are subjected to ambient-temperature chemical lysis and directly added to a closed-tube one-pot reaction containing RPA and CRISPR-Cas12a reagents. Following target amplification and sequence-specific recognition of MTB DNA, activated Cas12a cleaves reporter molecules, generating a signal that is visualized by lateral flow assay (LFA). The entire workflow enables rapid detection of MTB without nucleic acid extraction or complex laboratory instrumentation.

### 2.2. Control MTB DNA Templates

Genomic DNA from MTB strain H37Rv (ATCC 27294D; American Type Culture Collection, Manassas, VA) was used as the positive reference standard for assay optimization and analytical performance evaluation. The H37Rv strain is a well-characterized virulent laboratory reference strain widely used for molecular diagnostic development for detection of MTB [30]. DNA concentration and purity were verified upon receipt using spectrophotometric analysis and quantitative PCR (qPCR) targeting the *IS6110* insertion sequence. Stock DNA solutions were aliquoted and stored at −20°C to minimize freeze-thaw degradation. Serial tenfold dilutions of MTB genomic DNA were prepared in nuclease-free water to generate target concentrations ranging from 10^5^ to 1 copy per reaction for analytical sensitivity and limit-of-detection (LoD) studies. To determine assay performance in a clinically relevant matrix, defined quantities of MTB genomic DNA were spiked into MTB-negative sputum specimens prior to sample processing. Non-template controls (NTCs) containing nuclease-free water were included in all experiments. Assay specificity and potential cross-reactivity were evaluated using genomic DNA from clinically relevant non-tuberculous mycobacteria (NTM), including *Mycobacterium avium* (*M. avium*) and *Mycobacterium kansasii* (*M. kansasii*). Common respiratory bacterial pathogens, including *Staphylococcus aureus* (*S. aureus*) and *Streptococcus pneumoniae* (*S. pneumoniae*), were included in the specificity panel. These DNA preparations were also used for optimization of RPA primers and CRISPR guide RNAs, analytical sensitivity testing, and assay validation studies.

### 2.3. Ambient-Temperature Extract-Free Sample Preparation

To enable direct detection of MTB without conventional nucleic acid extraction, an ambient-temperature chemical lysis protocol was developed based on previously reported extraction-free strategies with modifications for sputum processing [20, 21, 30, 31]. Briefly, sputum specimens were mixed in a 1:1 volumetric ratio with a matrix-tolerant lysis buffer containing 1.5% (v/v) Triton X-100 (Sigma-Aldrich, St. Louis, MO), 1.0% (v/v) Tween-20 (Sigma-Aldrich), and 25 mM dithiothreitol (DTT) in an alkaline buffer (pH 8.5). The nonionic detergents were used to disrupt the lipid-rich mycolic acid-containing cell wall of MTB and facilitate release of genomic DNA, while DTT reduced sputum viscosity by breaking disulfide bonds within mucin proteins and helped minimize interference from endogenous proteins and nucleases. Following gentle mixing, samples were incubated at ambient temperature (22-25°C) for 5 minutes without heating, centrifugation, or mechanical disruption. After incubation, 2 μL of the resulting crude lysate was directly added to the one-pot RPA-CRISPR-Cas12a reaction mixture. No spin-column purification, magnetic-bead isolation, or additional nucleic acid processing steps were performed, thereby preserving a simplified sample-to-answer workflow suitable for point-of-care applications.

### 2.4. One-Pot RPA-CRISPR-Lateral Flow Assay (LFA)

The one-pot assay combined RPA, CRISPR-Cas12a detection, and LFA readout within a single reaction workflow [9–13, 31, 32]. Primers and CRISPR guide RNA (crRNA) were designed to target a conserved 140-bp region within the MTB-specific IS6110 insertion element. The primer and crRNA sequences were as follows: forward primer (RPA-F), 5′-GCTGAACGGCTGATGACCAAACTCGGCCTG-3′; reverse primer (RPA-R), 5′-GCTAGGGTGCTCGCCAGGTACTGATCATTG-3′; and crRNA, 5′-UAAUUUCUACUAAGUGUAGAUGCUUCGCCAGGTACTGATCA-3′. The 25 μL reaction mixture contained 1× isothermal reaction buffer (50 mM Tris-HCl, 10 mM (NH4)2SO4, 50 mM NaCl, pH 7.9), 400 nM of each primer, 250 nM LbCas12a (New England Biolabs, Ipswich, MA), 300 nM crRNA, 500 nM FAM- and biotin-labeled single-stranded DNA reporter probe, and 14 mM magnesium acetate. To minimize premature cleavage of amplification products, the Cas12a-crRNA ribonucleoprotein complex was initially retained separately during the first 10 min of amplification and subsequently introduced into the reaction by tube inversion. Reactions were incubated at ambient temperature. Upon recognition of the MTB target sequence, activated Cas12a cleaved the FAM- and biotin-labeled reporter probe through collateral trans-cleavage activity, generating products detectable by lateral flow analysis. Following completion of the one-pot RPA-CRISPR reaction, lateral flow strips (HybriDetect Universal LFA Kit, Milenia Biotec GmbH, Giessen, Germany) were inserted directly into the reaction tubes according to the manufacturer’s instructions. Positive samples generated both test and control bands, whereas negative samples generated only the control band (Figure 1). Test bands typically became visible within 15 min after strip insertion. Images of developed strips were captured using a standard smartphone camera for documentation and quantitative analysis. Test-line and control-line signal intensities were measured using ImageJ software (version 1.54, National Institutes of Health, Bethesda, MD). Relative signal intensity was expressed as the test-to-control (T/C) ratio and used for comparison among experimental groups[31, 33].

### 2.5. Clinical Specimens

Archived, de-identified sputum specimens were obtained at Jiangsu Province Hospital of Chinese Medicine. All procedures were conducted under institutional review board-approved protocol (IRB 253825), in accordance with ethical standards. The clinical validation cohort consisted of 100 specimens collected between October 2020 and December 2023, including 50 clinically confirmed pulmonary TB patients and 50 MTB-negative controls. Diagnosis of TB was established based on microbiological confirmation using GeneXpert MTB/RIF, culture, and/or qPCR testing according to standard clinical criteria [34, 35]. Available specimen information included TB status and corresponding molecular diagnostic results. Cohort characteristics are summarized in Table 1. To evaluate the extraction-free workflow, aliquots of each sputum specimen were processed using the ambient-temperature lysis protocol and directly analyzed by the one-pot RPA-CRISPR-Cas12a assay. In parallel, matched specimens underwent conventional nucleic acid extraction followed by qPCR analysis, and assay performance was compared with GeneXpert MTB/RIF results.

**Table 1:** Characteristics of the Archived Clinical Specimen Cohort.

| **Table 1: Characteristics of the Archived Clinical Specimen Cohort** |  |  |  |
| --- | --- | --- | --- |
| Characteristics | MTB-Positive Cases (n=50) | Healthy Controls (n=50) | P-value |
| Mean Age, years (SD) | 46.8 (11.5) | 44.2 (12.1) | 0.28 |
| Sex (Female/Male) | 18 / 32 | 21 / 29 | 0.54 |
| Specimen Matrix Type | Sputum (n = 50) | Sputum (n = 50) | N/A |
| Smear Microscopy Status | 38 Positive / 12 Negative | 50 Negative | < 0.001 |

### 2.6. Reference Testing (GeneXpert and qPCR)

Reference testing was performed using GeneXpert MTB/RIF (Cepheid, Sunnyvale, CA) and quantitative PCR (qPCR) assays according to established protocols [34–37]. GeneXpert served as the primary clinical comparator for evaluation of diagnostic performance. For qPCR analysis, genomic DNA was extracted from sputum specimens using validated laboratory procedures, and MTB was detected by amplification of the IS6110 insertion sequence using previously validated primer-probe sets [34–37]. MTB DNA concentrations were determined using standard curves generated from serial dilutions of quantified H37Rv genomic DNA and expressed as copies per reaction. To evaluate assay performance across a range of bacterial burdens, MTB-positive specimens were stratified according to qPCR Ct values into high-burden (Ct <25), moderate-burden (Ct 25–30), and low-burden (Ct >30) groups. qPCR served as the primary reference method for bacterial burden quantification, while GeneXpert MTB/RIF was used as a clinical comparator. Diagnostic sensitivity, specificity, predictive values, and overall concordance of the extraction-free RPA-CRISPR-Cas12a assay were then evaluated.

### 2.7. Statistical Analysis

The clinical validation cohort included 50 MTB-positive and 50 MTB-negative sputum specimens. A sample size of 50 MTB-positive and 50 MTB-negative specimens provided approximately 80% statistical power at a two-sided α level of 0.05 to evaluate assay sensitivity and specificity and to generate reasonably precise 95% confidence intervals for diagnostic performance estimates. Analytical sensitivity was determined using serial dilutions of quantified H37Rv genomic DNA, and the LoD was defined as the lowest concentration detected in ≥95% of replicate reactions. Probit regression analysis was used to estimate the concentration corresponding to a 95% probability of detection with 95% confidence intervals. Diagnostic performance, including sensitivity, specificity, and area under the receiver operating characteristic curve (AUC), was calculated with exact 95% confidence intervals. Agreement between the extraction-free RPA-CRISPR-Cas12a assay and reference methods was assessed using Cohen’s kappa coefficient. MTB-positive specimens were stratified by qPCR Ct values to evaluate assay performance across different bacterial burdens. Statistical analyses were performed using GraphPad Prism (v10.1), R (v4.3), and Python (v3.11), with P <0.05 considered statistically significant.

## 3. RESULTS

### 3.1. Analytical Specificity of the One-Pot RPA-CRISPR-Cas12a Assay

Species-specific RPA primers and a CRISPR-Cas12a crRNA targeting the highly conserved IS6110 insertion sequence were evaluated under optimized one-pot reaction conditions. Distinct lateral-flow test-band formation was observed exclusively in reactions containing MTB genomic DNA, whereas NTCs and non-target bacterial DNA controls generated only control bands and no visible test bands (Figure 2). Importantly, no cross-reactivity was observed with genomic DNA from clinically relevant NTM species, including *M. avium* and *M. kansasii*, or with common respiratory bacterial pathogens, including *S. aureus* and *S. pneumoniae*. These findings demonstrate the high analytical specificity of the IS6110-targeted one-pot RPA-CRISPR-Cas12a assay for detection of MTB.

**Figure 2.**
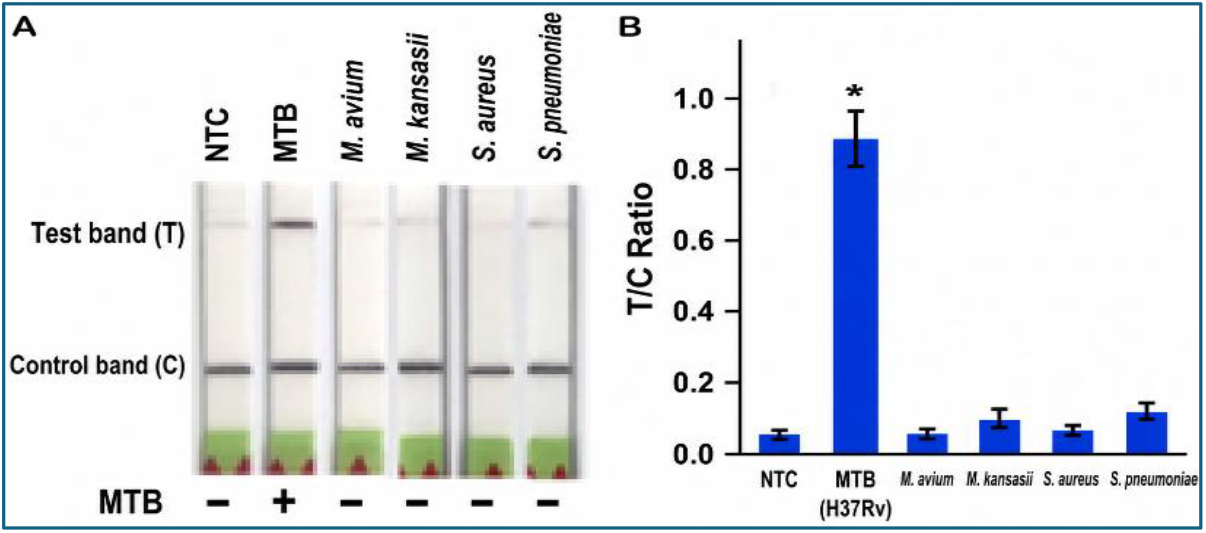
Analytical specificity of the one-pot RPA-CRISPR-Cas12a assay for detection of MTB. (A) Representative LFA results obtained using genomic DNA from M. tuberculosis H37Rv, a non-template control (NTC), non-tuberculous mycobacteria (*M. avium* and *M. kansasii*), and common respiratory bacterial pathogens (*S. aureus* and *S. pneumoniae*). A distinct test (T) band together with the control (C) band was observed only in reactions containing M. tuberculosis H37Rv DNA, whereas the NTC, NTM species, and non-target bacterial pathogens produced only the control band, demonstrating excellent analytical specificity without detectable cross-reactivity. (B) Quantitative analysis of LFA signals expressed as the test-to-control (T/C) ratio for the corresponding assays. Bar graphs represent the mean ± SD of three independent experiments. The MTB H37Rv sample exhibited a significantly higher T/C ratio than the NTC and all non-target organisms, which generated only background-level signals. *, *p* < 0.01.

### 3.2. Analytical Sensitivity of the One-Pot RPA-CRISPR-Cas12a Assay

The one-pot RPA-CRISPR-Cas12a assay was evaluated using ten-fold serial dilutions of quantified MTB H37Rv genomic DNA ranging from 10^5^ to 1 copy per reaction. A conventional two-step RPA-CRISPR workflow was performed in parallel using the same dilution series to enable direct comparison of analytical sensitivity and detection performance. Distinct lateral-flow test bands were observed across the dilution series, with test-band intensity decreasing proportionally with target copy number (Figure 3). Quantitative analysis of lateral-flow strips using ImageJ demonstrated a corresponding reduction in the T/C ratio as MTB DNA concentration decreased. The analytical LoD, defined as the lowest concentration detected in ≥95% of replicate reactions, was determined to be 10 copies per reaction based on three independent experiments. Probit regression analysis confirmed a ≥95% probability of detection within this concentration range (Figure 3), demonstrating that the one-pot assay achieved analytical sensitivity comparable to the conventional two-step assay while providing the advantages of a simplified closed-tube workflow.

**Figure 3.**
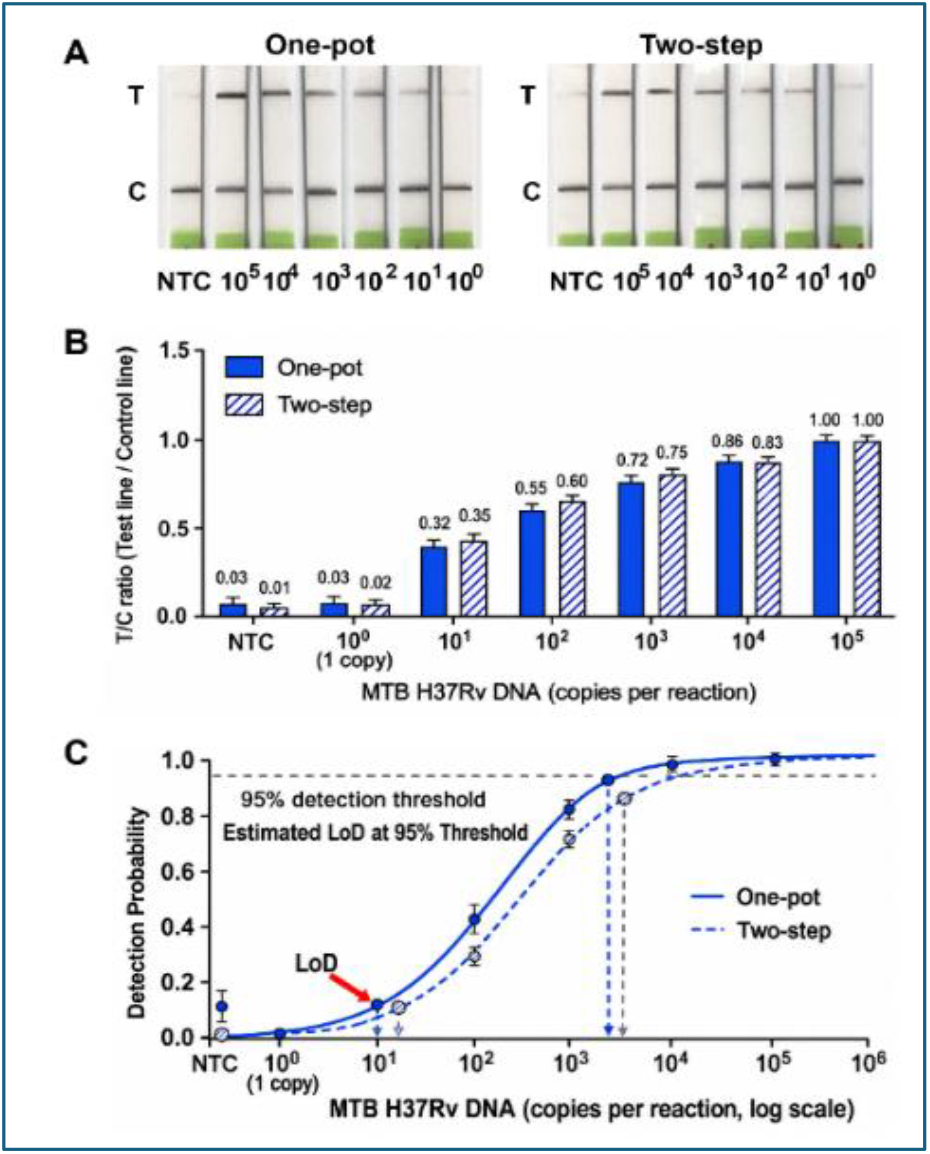
Analytical sensitivity of the one-pot RPA-CRISPR-Cas12a assay for detection of *Mycobacterium tuberculosis*. (A) Representative LFA strips obtained with the one-pot and conventional two-step RPA-CRISPR-Cas12a assays using ten-fold serial dilutions of *M. tuberculosis* H37Rv genomic DNA (10^5^–1 copy per reaction) together with a NTC. Test-band intensity decreased with decreasing DNA input, whereas no visible test band was detected in the NTC. (B) Quantification of LFA signals, expressed as the test-to-control (T/C) ratio, for the one-pot and two-step assays across the serial dilution series. Data are presented as mean ± SD from three independent experiments. (C) Probit regression analysis of detection probability as a function of MTB DNA concentration. The LoD at the 95% detection probability was 10 copies per reaction for the one-pot assay, comparable to that of the conventional two-step assay. The horizontal dashed line indicates the 95% detection threshold, and the vertical dashed lines indicate the corresponding estimated LoDs

### 3.3. Analytical Performance of the Extraction-Free One-Pot RPA-CRISPR-Cas12a Assay

To evaluate the feasibility of direct MTB detection without conventional nucleic acid extraction, serial dilutions of quantified H37Rv genomic DNA were spiked into MTB-negative sputum specimens and processed using the ambient-temperature extraction-free lysis protocol. The spiked specimens were divided into two portions for direct comparison of sample-processing workflows. One portion was analyzed using the extraction-free one-pot RPA-CRISPR-Cas12a assay, whereas the second portion underwent conventional DNA extraction followed by analysis using the one-pot RPA-CRISPR assay and reference qPCR.

Using purified DNA, the one-pot RPA-CRISPR assay demonstrated analytical sensitivity comparable to qPCR, with both methods achieving a LoD of 10 copies per reaction (Figure 4). In contrast, the extraction-free workflow using crude sputum lysates achieved a LoD of 100 copies per reaction, representing a one-log reduction in analytical sensitivity compared with purified-DNA testing. Nevertheless, distinct lateral-flow test bands were observed across clinically relevant MTB concentrations, and quantitative analysis of T/C ratios demonstrated robust target detection. Furthermore, direct introduction of crude sputum lysates did not substantially inhibit RPA amplification or CRISPR-Cas12a activity, indicating that the extraction-free workflow was compatible with downstream molecular detection. No false-positive signals were observed among negative controls, and analytical specificity remained 100% across all assay formats tested. These findings indicate that the extraction-free workflow preserves high analytical specificity while substantially simplifying sample processing by eliminating nucleic acid purification. The ability to directly detect MTB from minimally processed sputum supports the feasibility of a streamlined sample-to-answer molecular diagnostic platform for POC applications.

**Figure 4.**
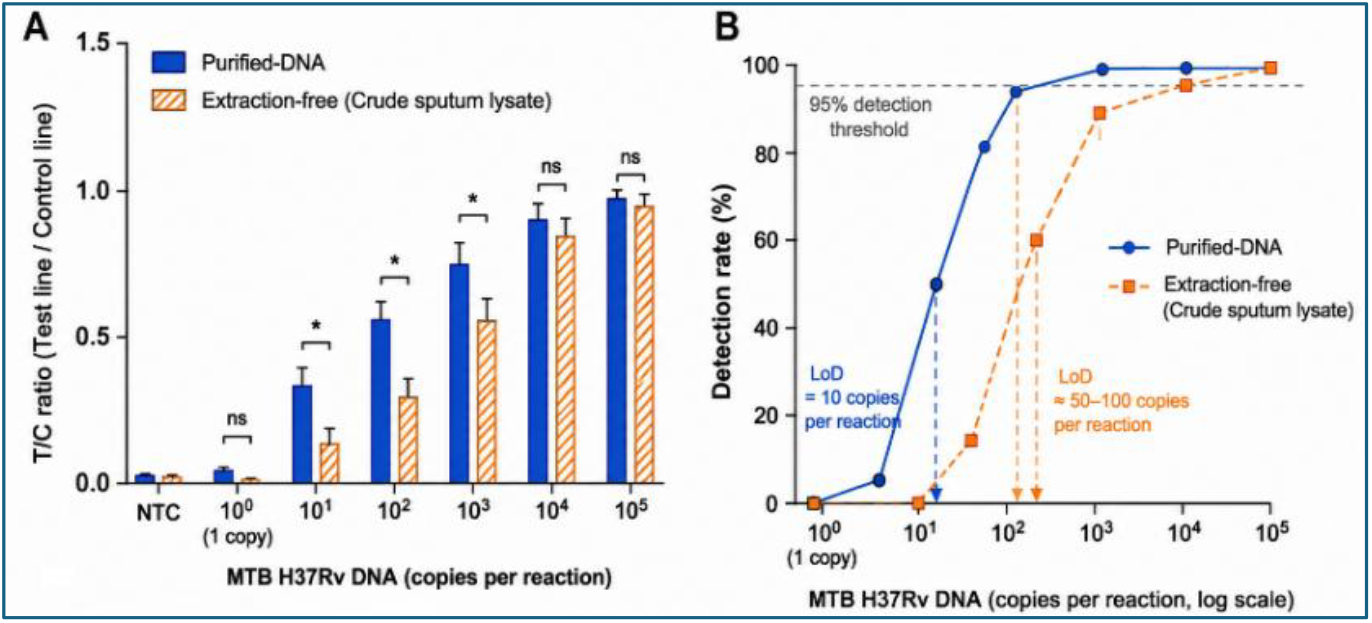
Analytical performance of the extraction-free one-pot RPA-CRISPR-Cas12a assay compared with the purified-DNA workflow. (A) Comparison of LFA signal intensity, expressed as the T/C ratio, between the purified-DNA and extraction-free (crude sputum lysate) workflows using ten-fold serial dilutions of MTB H37Rv genomic DNA. Data are presented as mean ± SD from three independent experiments. Statistical significance between the two workflows at each DNA concentration is indicated (*, p < 0.05; ns, not significant). (B) Detection probability of the purified-DNA and extraction-free workflows across serial MTB DNA concentrations. The LoD was defined as the lowest DNA concentration detected in ≥95% of replicate reactions. The purified-DNA workflow achieved an estimated LoD of 10 copies per reaction, whereas the extraction-free workflow achieved an estimated LoD of 100 copies per reaction.

### 3.4. Diagnostic Performance of the Extraction-Free One-Pot RPA-CRISPR-Cas12a Assay in Clinical Specimens

Among the 50 MTB-positive specimens, 22 (44.0%) exhibited high bacterial burden (qPCR Ct <25), 18 (36.0%) moderate burden (Ct 25–30), and 10 (20.0%) low burden (Ct >30). The extraction-free one-pot RPA-CRISPR-Cas12a assay correctly identified 48 of 50 MTB-positive specimens and 49 of 50 MTB-negative specimens. Two false-negative results occurred among low-burden specimens, whereas one false-positive result was observed among MTB-negative controls. ROC analysis demonstrated excellent diagnostic performance, with an area under the curve (AUC) of 0.982 (95% CI: 0.954–1.000; P < 0.001) (Figure 5). Overall diagnostic sensitivity and specificity were 96.0% and 98.0%, respectively (Table 3).

**Figure 5.**
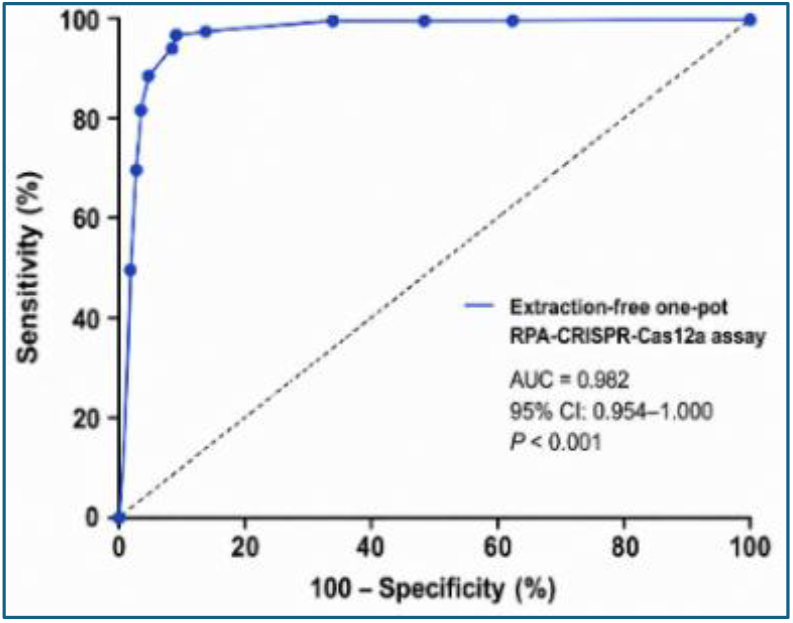
Diagnostic performance of the extraction-free one-pot RPA-CRISPR-Cas12a assay in sputum specimens. ROC curve demonstrating the clinical diagnostic performance of the extraction-free one-pot RPA-CRISPR-Cas12a assay for detection of MTB in sputum specimens. The assay achieved an area under the curve (AUC) of 0.982 (95% CI: 0.954– 1.000; *P* < 0.001), indicating excellent discrimination between MTB-positive and MTB-negative specimens. The simplified extraction-free workflow maintains high diagnostic accuracy while minimizing sample processing and supporting rapid molecular detection in clinical settings.

Agreement with reference methods (GeneXpert MTB/RIF and qPCR) was high, yielding an overall concordance of 97.0% (97/100) and a Cohen’s κ coefficient of 0.94, indicating near-perfect agreement (Table 2). To assess the impact of bacterial burden, LFA signal intensity, expressed as the T/C ratio, was analyzed in relation to qPCR Ct values. A significant inverse correlation was observed between Ct value and T/C ratio (P < 0.05), indicating stronger assay signals in specimens with higher MTB DNA concentrations (Figure 6A). Logistic regression analysis further showed that the probability of detection decreased as Ct values increased (P < 0.05) (Figure 6B). The assay detected all high- and moderate-burden specimens (100% sensitivity), whereas sensitivity decreased to 80% (8/10) among low-burden specimens (Table 3). Collectively, these findings demonstrate that the extraction-free one-pot RPA-CRISPR-Cas12a assay provides accurate MTB detection directly from minimally processed sputum while eliminating conventional nucleic acid extraction, supporting its potential use as a simplified POC diagnostic platform for tuberculosis.

**Table 2.** Diagnostic Performance of the Extraction-Free One-Pot RPA-CRISPR-Cas12a Assay for Detection of MTB in Clinical Sputum Specimens.

| Metric | Value | 95% CI |
| --- | --- | --- |
| Sensitivity | 96.0% | 86.3-99.5 |
| Specificity | 98.0% | 89.4-99.9 |
| Cohen's $\kappa$ | 0.94 | |
| AUC | 0.982 | 0.954-1.000 |

**Table 3.**
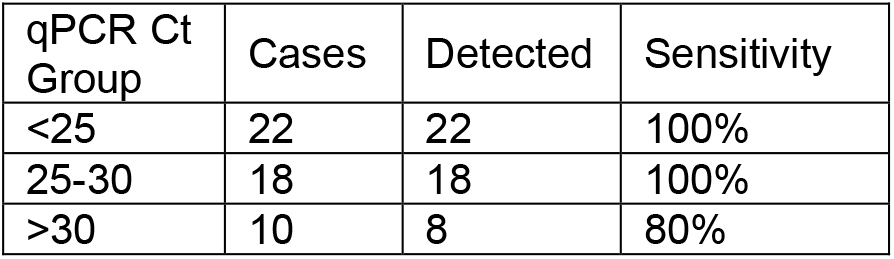
Clinical Sensitivity of the Extraction-Free One-Pot RPA-CRISPR-Cas12a Assay Stratified by MTBBurden Determined by qPCR Ct Values.

**Figure 6.**
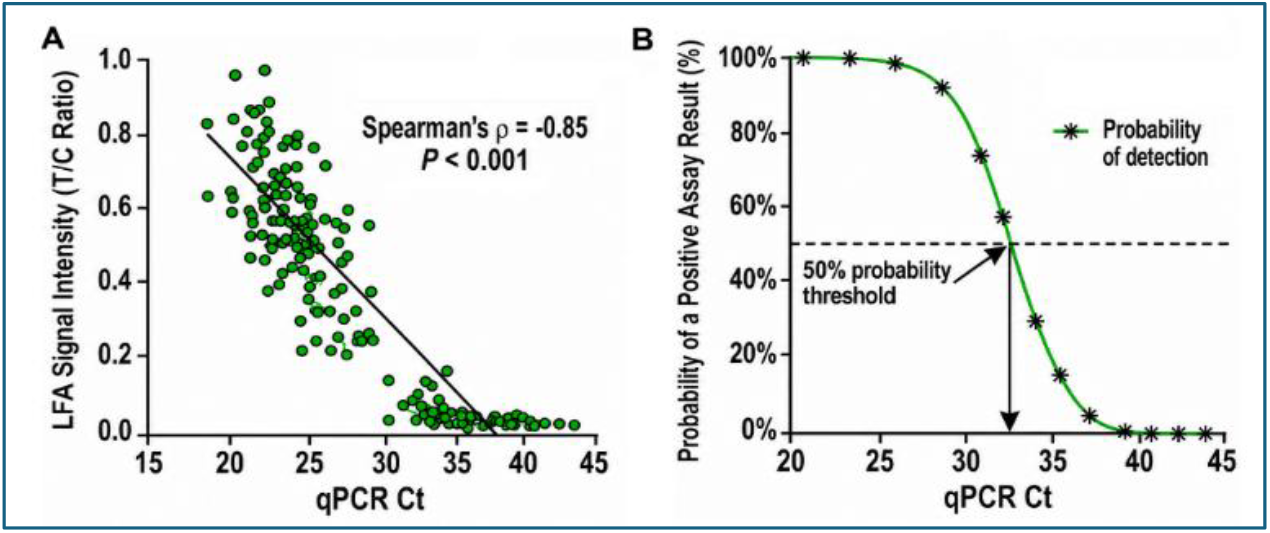
Relationship between bacterial burden and assay performance. (A) Correlation between qPCR Ct values and LFA signal intensity, expressed as the T/C ratio, in MTB-positive clinical sputum specimens. Each point represents one clinical specimen. A significant inverse correlation was observed (Spearman’s ρ = −0.85, P < 0.001). (B) Logistic regression analysis showing the probability of a positive extraction-free one-pot RPA-CRISPR-Cas12a assay result as a function of qPCR Ct value. Detection probability decreased with increasing Ct values, indicating reduced assay sensitivity at lower bacterial burdens. The dashed line indicates the 50% probability threshold.

## 4. DISCUSSION

Molecular assays, including GeneXpert and conventional PCR, have substantially improved the sensitivity, specificity, and turnaround time of TB diagnosis compared with smear microscopy [6]. However, these methods require specialized instruments, stable electricity, trained personnel, and relatively high costs, limiting their implementation in resource-limited and point-of-care settings [6]. Several CRISPR-based assays have been developed for TB diagnosis using diverse CRISPR effectors, including Cas12a, Cas12b, Cas12f, and Cas13a [17–29]. As summarized in Table 4, these platforms have progressively improved the analytical performance of CRISPR diagnostics by incorporating either extraction-free processing or one-pot amplification. However, most still require nucleic acid extraction, multi-step sample preparation, laboratory instrumentation, heating, or combinations.

**Table 4.** Comparison of Representative CRISPR-Based MTB Detection Platforms Published Between 2019 and 2025.

| **Study** | **CRISPR System** | **Direct Sputum** | **Extraction -Free** | **One-Pot** | **Ambient-Temperature Processing** |
| --- | --- | --- | --- | --- | --- |
| Ai et al., [17] | Cas12a | No | No | No | No |
| Ren et al., [28] | Cas13a | No | No | No | No |
| Tram et al., [29] | CRISPR-based NGS | Yes | Partial | No | No |
| Peng et al., [24] | Cas12b + CPA | Yes | No | Yes | No |
| Deng et al., [27] | Cas12f1_ge4.1 + RPA | No | No | No | No |
| Bell et al., [21] | Cas13a + Cas12a QC | Yes | Yes | Partial | Not fully ambient-temperature |
| Present Study | Cas12a + RPA | Yes | Yes | Yes | Yes |

Compared with these previously reported platforms, the assay developed in this study uniquely integrates four key workflow innovations into a single molecular diagnostic platform: (i) direct analysis of minimally processed sputum, (ii) extraction-free sample preparation, (iii) one-pot closed-tube RPA-CRISPR-Cas12a detection, and (iv) entirely ambient-temperature sample processing. To our knowledge, none of the representative CRISPR-based MTB assays reported to date incorporate all four features within a single workflow (Table 4). By eliminating nucleic acid extraction, heating, centrifugation, and mechanical disruption, the assay substantially simplifies molecular testing while minimizing contamination risk associated with conventional multi-step workflows. Despite this simplified workflow, the assay maintained robust analytical and clinical performance. Using crude sputum lysates, it achieved an analytical limit of detection of 100 copies per reaction within 30 minutes, indicating that ambient-temperature chemical lysis is fully compatible with downstream RPA amplification and CRISPR-Cas12a detection. Furthermore, direct comparison with purified-DNA workflows demonstrated only a modest reduction in analytical sensitivity while preserving high diagnostic accuracy in clinical specimens. These findings suggest that conventional nucleic acid purification may not be essential for accurate CRISPR-based MTB detection. An additional strength of this study is the comprehensive evaluation of the extraction-free workflow. Unlike most previous reports [17–29], which focused primarily on analytical proof-of-concept, we directly compared extraction-free and purified-DNA workflows, assessed analytical specificity and sensitivity, and validated diagnostic performance in a clinical sputum cohort. Together, these findings provide practical evidence supporting implementation of an integrated, sample-to-answer CRISPR platform for decentralized and POC molecular diagnosis of tuberculosis, particularly in resource-limited settings.

Several limitations should be acknowledged. First, the study was conducted using archived clinical specimens under controlled laboratory conditions. Prospective validation using freshly collected specimens is needed to confirm assay performance in real-world clinical settings. Second, the sample size was relatively modest and derived from a limited number of collection sites. Additional multicenter studies involving geographically diverse populations will be necessary to further establish clinical utility. Third, although the assay demonstrated high diagnostic accuracy, additional studies are needed to evaluate performance in smear-negative, paucibacillary, pediatric, and extrapulmonary tuberculosis cases. Finally, long-term stability testing of assay reagents under varying environmental conditions has not yet been completed. Future studies will focus on further optimization of assay robustness, evaluation in larger prospective cohorts, and expansion of the platform to additional specimen types.

## 5. CONCLUSIONS

This study demonstrates the feasibility of an extraction-free one-pot CRISPR-Cas12a assay for rapid detection of MTB directly from minimally processed sputum specimens. The platform achieved high analytical sensitivity while eliminating conventional nucleic acid extraction procedures. By simplifying specimen processing and integrating amplification and detection within a single closed-tube workflow, this approach may facilitate the implementation of POC TB diagnostics and decentralized molecular testing in resource-limited settings.

## Ethics Approval and Consent to Participate

The study protocol (IRB 253825) was reviewed and approved by Jiangsu Province Hospital of Chinese Medicine. All procedures were conducted under institutional review board-approved protocol, following the ethical standards. Written informed consent was obtained from all participants prior to sample collection and study enrollment.

## CRediT authorship contribution statement

Jipei Liao: Conceptualization, Methodology, Investigation, Data curation, Formal analysis, Visualization, Writing – original draft. Yun Su: Investigation, Resources, Sample collection, Validation, Writing – review & editing. Feng Jiang: Conceptualization, Supervision, Project administration, Funding acquisition, Writing – review & editing.

## Acknowledgements

The authors wish to thank the Biostatistics Shared Service at the University of Maryland Marlene and Stewart Greenebaum Cancer Center for their invaluable contribution in conducting the statistical analysis for this study.

## Data availability

The data supporting the findings of this study are available within the article and its supplementary materials.

## MANUSCRIPT FIGURES AND TABLES

1. **Figure 1. Extraction-Free One-Pot RPA-CRISPR-Cas12a Assay Workflow**. Schematic illustration of the ambient-temperature sputum lysis protocol followed by direct introduction of crude sputum lysate into a closed-tube one-pot RPA-CRISPR-Cas12a reaction and lateral-flow assay (LFA) readout, enabling rapid sample-to-answer detection of *Mycobacterium tuberculosis* (MTB) without nucleic acid extraction.
2. **Figure 2. Analytical Specificity of the Extraction-Free One-Pot RPA-CRISPR-Cas12a Assay**. Representative LFA strips and quantitative analysis of test-to-control (T/C) ratios demonstrating specific detection of MTB DNA without cross-reactivity to non-tuberculous mycobacteria or common respiratory bacterial pathogens.
3. **Figure 3. Analytical Sensitivity and Comparison of One-Pot and Conventional Two-Step RPA-CRISPR-Cas12a Assays**. Representative LFA strips, quantitative T/C ratios across serial MTB genomic DNA dilutions, and probit regression analysis comparing the analytical sensitivity of the one-pot and conventional two-step assays, demonstrating comparable limits of detection of 10 copies per reaction.
4. **Figure 4. Analytical Performance of the Extraction-Free Workflow Compared with Purified-DNA Testing**. Comparison of purified-DNA and extraction-free sputum workflows using quantitative LFA T/C ratios and detection probability analysis across serial MTB DNA concentrations, demonstrating preserved analytical performance with a modest reduction in sensitivity following extraction-free sample processing.
5. **Figure 5. Diagnostic Performance of the Extraction-Free One-Pot RPA-CRISPR-Cas12a Assay in Clinical Sputum Specimens**. Receiver operating characteristic (ROC) curve demonstrating excellent diagnostic accuracy for detection of MTB, with an area under the curve (AUC) of 0.982.
6. **Figure 6. Relationship Between Bacterial Burden and Assay Performance**. Correlation between qPCR Ct values and LFA signal intensity (T/C ratio), together with logistic regression analysis of detection probability across different bacterial burdens, demonstrating reduced assay sensitivity with decreasing MTB DNA concentration.

1. **Table 1. Characteristics of the Clinical Sputum Specimen Cohort.** Demographic and clinical characteristics of the archived sputum specimens used for clinical validation, including age, sex, specimen type, and smear microscopy status for MTB-positive patients and MTB-negative controls.
2. **Table 2. Diagnostic Performance of the Extraction-Free One-Pot RPA-CRISPR-Cas12a Assay for Detection of *Mycobacterium tuberculosis* in Clinical Sputum Specimens**. Summary of diagnostic sensitivity, specificity, area under the receiver operating characteristic curve (AUC), and Cohen’s κ coefficient with corresponding 95% confidence intervals where applicable.
3. **Table 3. Diagnostic Performance of the Extraction-Free One-Pot RPA-CRISPR-Cas12a Assay Stratified by Bacterial Burden**. Detection rates and diagnostic sensitivity of the assay across clinical specimens grouped by qPCR cycle threshold (Ct) values representing high-, moderate-, and low-bacterial-burden tuberculosis.
4. **Table 4. Comparison of Representative CRISPR-Based *Mycobacterium tuberculosis* Detection Platforms Published Between 2019 and 2025**. Comparison of representative CRISPR-based MTB diagnostic platforms with respect to CRISPR system, direct sputum testing, extraction-free processing, one-pot assay format, and ambient-temperature sample processing.

